# Theraputic Effects of Brain-Computer Interface on Motor Recovery of Stroke Patients: A Meta-analysis

**DOI:** 10.1101/2023.04.11.23288439

**Authors:** Zhiwei Guo, Qiang Gao, Yi Jiang, Hanhong Jiang, Ning Jiang

## Abstract

**Background:** Previous clinical studies have demonstrated the effects of brain-computer interface (BCI) on the motor recovery of stroke patients. The aim of this study was to evaluate the therapeutic effects of BCI on improving motor functions of stroke patients.

**Methods:** We conducted a meta-analysis on randomized controlled trials (RCTs) on BCI training for post-stroke motor rehabilitation. Relevant publications were identified from the databases of PubMed, Embase, ScienceDirect, and Cochrane Library. The standardized mean difference (SMD) with 95% confidence intervals (CI) were calculated as the pooled effect size of the motor outcome.

**Results:** Thirty-five of the 43 candidate articles involving 749 participants were included in this meta-analysis. Overall, both the significant immediate effect size of 0.53 and long-lasting effect size of 0.26 were found for motor outcome measured by Fugl-Meyer Assessment. A further subgroup-analysis observed larger therapeutic effects on lower-limb than upper-limb. A subgroup-analysis also indicated that stroke patients may gain better functional outcome in the subacute phase than in the chronic phase. Superior effect of BCI training was also detected for distal function of upper-limb over proximal function. BCI training combined with functional electrical stimulation (FES) was more effective than BCI combined with robot. No significant effect was found in other combined interventional methods, especially the use of transcranial direct current stimulation, which cannot potentiate the effects of BCI training. In addition, subgroup-analysis also indicated a greater effect for longer durations of intervention. And the dosage between 15 min and 180 min was found to be optimal.

**Conclusions:** BCI has significant immediate and long-lasting effects on improving motor function of both upper-limb and lower-limb of stroke patients. Superior therapeutic could be delivered to patients in the subacute phase and clearer benefits are evident in distal functions of upper extremity. When combined with FES, BCI seems to be more effective than when combined with robot and other external devices. Longer durations of intervention could provide better effects. But bigger is not always better for weekly dosage.

## Introduction

Stroke is one of the leading causes of disability and death among adults worldwide [1]. More than 80% stroke survivors suffer acute motor dysfunction and more than 50% stroke survivors suffer from permanent motor impairment [2]. Motor deficit of either upper- or lower-limb could vastly result in reduced self-care ability in activities of daily living and seriously decreased quality of life [3]. Therefore, timely and effective rehabilitation of the motor function for stroke patients are needed drastically and very important.

Various types of post-stroke rehabilitation have been applied to facilitate motor recovery, including conventional therapy, electrical stimulation (e.g., noninvasive brain stimulation, vague nerve stimulation, or functional electric stimulation (FES)), and robot-assisted therapy [4–7]. However, these approaches are not suitable for all stroke patients due to variability across patient population. Recently, stroke rehabilitation based on neurofeedback and the mirror neuron system (e.g., motor imagery (MI), action observation (AO)) have been proposed as potentially alternative or complement methods [8–11]. Both methods can be realized as part of a brain-computer interface (BCI) system for stroke rehabilitation, through which the patient’s movement volitions can be decoded in real-time, and in turn triggering contingent sensory feedback [12–14]. With stroke patient with significant paralysis, BCIs decodes the patient’s volition through various technologies such as electroencephalography (EEG), functional near-infrared spectroscopy (fNIRS), or magnetoencephalography (MEG), all of which are non-invasive [15]. Upon the detection of patient’s movement volition, the feedback would be triggered and subsequently delivered by various forms that reproduce some physiological characteristics of the intended movement. These feedback methods include 1) abstract forms (e.g. moving cursor on a computer screen), 2) visual representation of the virtual limbs, and 3) somatosensory representations delivered through mechanical or electrical stimulation [16].

Therefore, BCI has been investigated in the clinical application as an assistive or rehabilitative technology to restore lost functions or facilitate motor recovery of stroke patients. A large body of literature investigated the effectiveness of BCI in post-stroke motor rehabilitation with first study published as early as 2008 [17]. In the last few years, several meta-analyses have shown that BCI does have significant effects on the improvement of hemiparetic upper-limb of stroke patients than conventional therapy [16–18]. Furthermore, comparisons of the effectiveness among different devices combined with BCI indicated that the BCI-FES combination may be the preferred method for functional recovery over other types of neural feedback [18, 19]. Although, the comparison of the effects between different phases (subacute vs chronic phase) of stroke recovery were conducted in two recent meta-analyses [16, 19], the results were inconsistent. Furthermore, the clinical efficacy on functions of the lower-limb, proximal and distal function of upper-limb, and optimal parameters, such as therapy duration, therapy frequency and of BCI training, are all still unknown. In order to provide answers to these questions, we present an updated meta-analysis with the following objectives: (1) to investigate potential differential effects of BCIs on upper-limb and lower-limb, (2) to assess the relative effects of BCIs on proximal and distal function of upper-limb, (3) to explore the optimal parameters such as treatment duration, frequency of BCI training for motor recovery of stroke patients with motor dysfunction, and (4) to undertake an update meta-analysis of all studies comparing the efficacy of BCIs when combined with different kinds of external devices (e.g., robot, FES, tDCS, and visual feedback).

## Methods

This study is registered through International Prospective Register of Systematic Reviews (PROSPERO, https://www.crd.york.ac.uk/PROSPERO/, ID: CRD42023410371) and is reported following the PRISMA reporting guideline.

### Search Strategy and Selection Criteria

To identify relevant studies, a systematic computerized literature searches were performed in three databases (PubMed, Embase, ScienceDirect, and Cochrane Library) with the following keywords: “stroke” and (“brain computer interface” or “BCI” or “brain machine interface” or “BMI”). The searches were conducted from the inception of each database to March, 2023 and limited to human studies published in English.

Studies were included in the current meta-analysis if they met the following criteria: (1) they were randomized controlled trials to investigate the effect of BCI/BMI on the upper/lower extremity motor functional recovery; (2) the subjects were adult stroke patients; (3) the outcome measures were reported with continuous scales that evaluated the motor function of the affected limb; (4) the outcome assessments could be expressed as mean ± standard deviation (SD); (5) the characteristics of the brain signal from the BCI system was used to decode the subjective motor intention of the patient in order to provide feedback for motor rehabilitation. Two investigators independently searched and screened the literature based on the titles and abstracts after exclusion of duplicated studies. Then, the full text of the remaining articles that appeared to be relevant were further reviewed carefully to evaluate the final eligibility. Any discrepancy was resolved by discussion with a third reviewer to reach an agreement.

### Quality Assessment

We evaluated the quality and the risk of bias of the included studies by using the Cochrane Collaboration’s risk of bias tool, containing seven aspects: (1) random sequence generation; (2) allocation concealment; (3) blinding of participants and personnel; (4) blinding of outcome assessment; (5) incomplete outcome data; (4) selective reporting and (7) other issues. These aspects could be used to assess the selection bias, performance bias, attrition bias, detection bias and reporting bias of each study. The judgement of bias of each aspect could be expressed simply as “Low risk”, “Unclear risk”, and “High risk” and selected in the tool. “Unclear risk” means that there is insufficient information to make this judgement or if the item is not relevant to the study.

### Data Extraction

The relevant data from each included study were extracted by two authors independently. The following information were extracted: year of publication, name of the first author, sample size, mean age, stroke duration, intervention protocol, type of control group, outcome measures, intervention and follow-up duration. For each outcome measure on motor function, the mean and SD of the pre- and post-intervention score changes were extracted, for both the experimental and control group. When the mean and SD of the change scores were not provided in the article, they were calculated with the reported pre- and post-data by the following formulae based on the principles of the Cochrane Handbook for systematic Reviews of Intervention (Higgins, 2011):

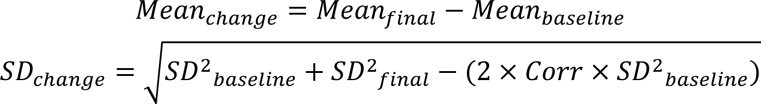

where Corr would be imputed from the studies which provided the baseline, final, and change standard deviation: 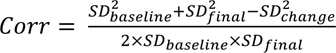. Otherwise, an average value of 0.8 was used following the recommendation in the Cochrane Handbook for Systematic Reviews of Interventions.

When the mean and standard error (SE) of the outcome measure was provided in the study, we transferred the SE value to SD with the following formula: SD = 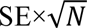. N represents the sample size. When the results of the outcome measure were not available and were reported as a graph, the data can be extracted and calculated by using the digitizing software of GetData Graph Digitizer 2.26 (http://getdata-graph-digitizer.com/).

### Statistical Analysis

The data from the motor outcome measures were used in the quantitative syntheses analyses which were performed with Review Manager Software version 5.4 (Cochrane Collaboration, Oxford, England). Considering the variety of the motor outcome measures used in the included studies, the standard mean difference (SMD) with 95% confidence intervals (CI) was calculated to estimate the pooled effect size after combining the weighted effect size of each study. To avert overestimation bias of the studies with small sample sizes, the weights were adjusted according to the sample size of respective studies automatically by the software. The heterogeneity was evaluated with the Cochrane’s Q test and Higgins’ I^2^ index. When the I^2^ value was < 50%, a fixed-effects model was applied in the syntheses analysis, which was considered acceptable homogeneity. In contrast, the random-effects model was used when I^2^ was ≥ 50% to accommodate the significant heterogeneity. Subsequent subgroup analysis and sensitivity analysis were also conducted to explore the source of heterogeneity in these cases. The funnel plots were used to detect the potential risk of publication bias. No significant publication bias was considered when an approximately symmetrical inverse funnel plot was obtained and a majority of the studies were located at the superior part of the plot. The asymmetry of the funnel plot was further quantified by using Egger’s linear regression. A two-sided *p* < 0.05 was regarded as statistically significant.

## Results

### Study selection

The screening and identification process resulted in a total of 43 full-texts articles. A further eligibility assessment by full-text review [13, 14, 20–60] excluded seven studies for with the following reasons: (1) two studies were excluded due to duplicate data from the same research group [53, 55], (2) mean and SD of the outcome data were not provided and cannot be calculated in three studies [54, 58, 59], (3) the EEG data of BCI were not used for motor rehabilitation in two studies [56, 57], and (4) one study did not provide complete data for meta-analysis [60]. Finally, 35 RCT studies were included in the quantitative synthesis [13, 14, 20–52]. A flow diagram of the study selection is presented in **Figure 1**.

**Figure 1.**
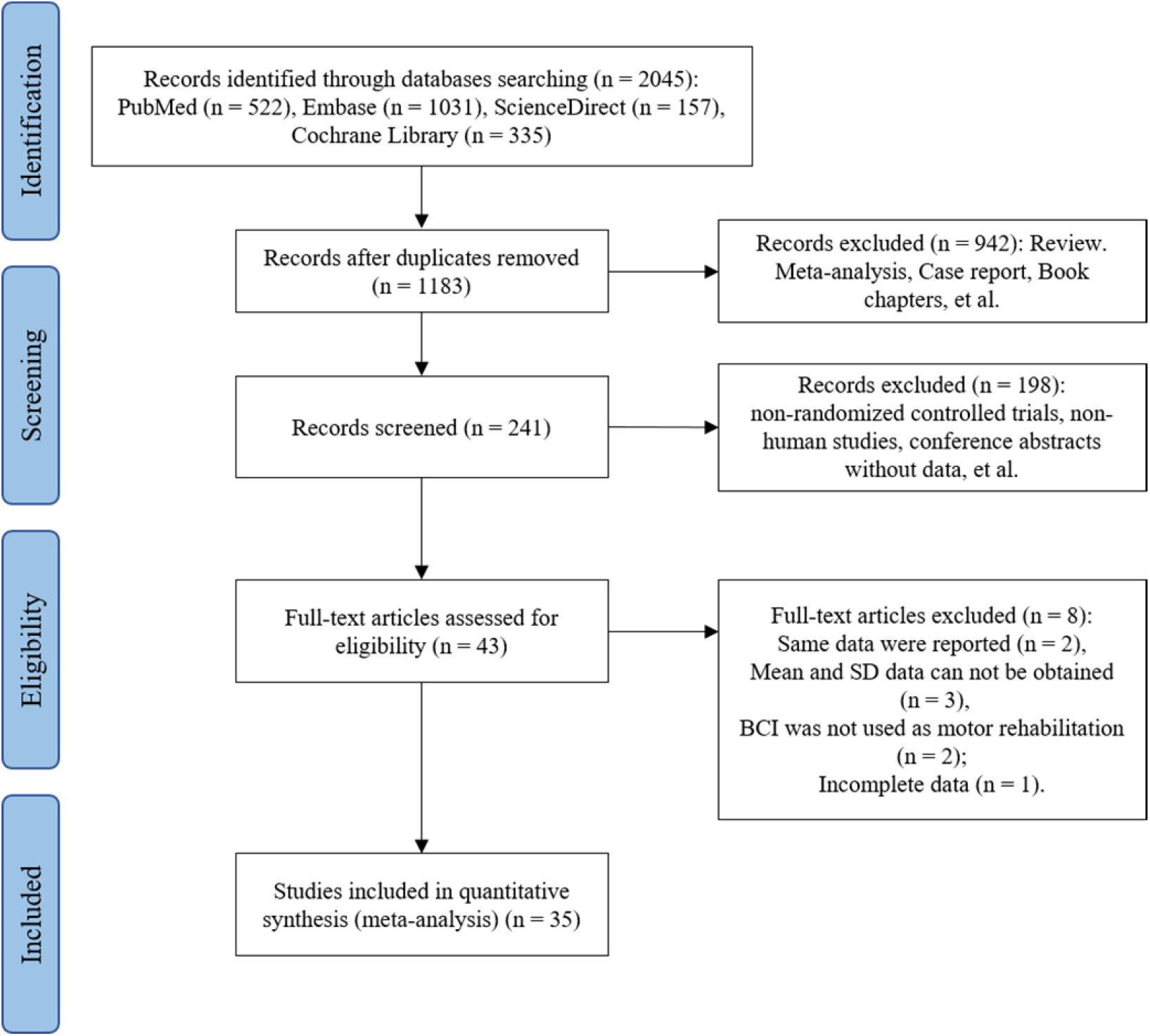
Flow diagram of study selection

### Characteristic of the included studies

The main characteristics of the included studies in the meta-analysis are shown in **Table 1**. Twenty-eight studies targeted stroke patients with upper-limb disability, seven studies targeted the lower-limb disability. Fourteen studies recruited stroke patients in the subacute stage (7 days to 6 months since stroke onset), 21 studies focused on stroke patients in the chronic stage (> 6 months since stroke onset), whereas one study recruited both subacute and chronic stroke patients. Regarding the BCI interventions, the motor intention signals detected from EEG or near-infrared spectroscopy (NIRS) were used to trigger external devices (e.g., robot, FES system, or visual feedback) for motor rehabilitation in all of the included studies. Event-related desynchronization (ERD)/event-related synchronization (ERS), steady-state visually evoked potentials (SSVEP), movement-related cortical potentials (MRCP) measured by EEG and task-related regional hemodynamic changes detected by NIRS were commonly used as the features of movement intention and decoded in real-time. Four of the studies also explored the effects of tDCS in facilitating the effects of BCIs on the improvement of motor recovery in the upper extremity. Besides, various outcome measures including Fugl-Meyer Assessment (FMA), Action Research Arm Test (ARAT), Wolf Motor Function Test (WMFT), Modified Barthel Index (MBI), and Motor Activity Log (MAL) were used in the selected studies to assess the immediate and long-term clinical effects of BCI on different aspects of motor function of stroke patients. The items of FMA scales of upper-limb consist of two categories, which are used to assess the proximal and distal function. The proximal part includes functions of shoulder, elbow, and arm, whereas the distal part includes functions for hand, wrist, and finger.

**Table 1.** Characteristics of the included studies.

|  | Year | Authors | No of participants (Exp/Cont) | Age (years) (Mean±SD) | Stroke Duration (months) (Mean±SD) | BCI interventions methods | Control group | Dosage of BCI | Outcome measures | Limbs |
| --- | --- | --- | --- | --- | --- | --- | --- | --- | --- | --- |
| 1 | 2014 | Ang et al | 21(6/8/7) | 54.2±12.4 | 12.83±4.39 | BCI to drive haptic knob | (1) Haptic knob<br>(2) Standard arm therapy | 90mins/day, 3 d/w, 6w(27h) | FMA | upper-limb |
| 2 | 2015 | Ang et al | 25(11/14) | 51.4±11.5 | 9.91±7.96 | BCI to drive MIT-Manus robot | MIT-Manus | 90mins/day, 3days/w, 4w(18h) | FMA | upper-limb |
| 3 | 2021 | Cantillo-Negrete et al | 10 | 59.9±12.8 | 4.67±2.77 | BCI to drive robotic orthosis | Conventional therapy. | 30-40mins/day, 12days(6-8h) | FMA, ARAT | upper-limb |
| 4 | 2020 | Chen et al | 14(7/7) | 41.6±12.0/<br>52.0±11.1 | 3.1±1.7/<br>3.9±1.5 | BCI to drive exoskeleton | Traditional therapy | 16.5mins/day, 3days/w, 4w | FMA | upper-limb |
| 5 | 2020 | Cheng et al | 10(5/5) | 62.4±4.7/<br>61.4±4.5 | 15.89±10.07/<br>29.67±8.57 | BCI to drive soft robotic glove | Soft robotic glove | 90mins/day, 3 d/w, 6w(27h) | FMA, ARAT | upper-limb |
| 6 | 2022 | Guo et al | 30(10/10/10) | 60.2±9.3/<br>56.9±6.1/<br>53.5±8.3 | 12.5±7.1/<br>11.7±5.4/<br>10.9±7.9 | SSVEP-BCI to drive soft robotic glove | 1. Soft robotic glove<br>2. Conventional therapy | 60mins/days, 5days/w, 2w(10h) | FMA, WMFT, MAS | upper-limb |
| 7 | 2015 | Curado et al | 30(16/14) | 46.56±13.44/<br>49.86±13.01 | 5.63±3.86/<br>5.50±6.06 | BMI to drive robotic orthosis | Sham BMI | 60mins/day, 5days/w, 4w(20h) | FMA | upper-limb |
| 8 | 2022 | Li et al | 24(12/12) | 43.8±14.7/<br>55.0±12.2 | Chronic and subacute stroke | BCI to drive exoskeleton hand robot | Conventional treatment | 60mins/day, 5days/w, 2w(10h) | FMA, ADL, WMFT, MBI | upper-limb |
| 9 | 2013 | Ramos et al | 32(16/16) | 49.3±12.5/<br>50.3±12.2 | 66±45/<br>71±72 | BCI to drive robotic orthosis | Sham BCI | 60 mins/day, 5days/w, 4w(20h) | cFMA, GAS, MAL, Ashworth | upper-limb |
| 10 | 2019 | Ramos et al | 28(16/12) | 49.3±12.5/<br>50.3±12.2 | Chronic stroke | BMI to drive robotic orthosis | Sham BMI | 60mins/days, 5days/w, 4w(20h) | FMA, MAL, GAS | upper-limb |
| 11 | 2019 | Ray et al | 30(16/14) | 49.8±12.4 | 68.5±58.5 | BCI to drive robotic orthosis | Sham BCI | 40mins/session, 17 sessions, 6w | FMA | upper-limb |
| 12 | 2019 | Wang et al | 24(13/11) | 54±9/<br>54±9 | 44.76±45.72/<br>42.6±24.24 | AOT-BCI to drive<br>robotic orthosis | Robot | 60mins/d, 5ays d/w,<br>4w(20 h) | FMA, ARAT,<br>WMFT | upper-limb |
| 13 | 2020 | Wu et al | 23(14/11) | 62.93±10.56/<br>64.82±7.22 | 2.11±0.30/ | BCI to drive<br>exoskeleton | Conventional<br>Treatment | 60mins/days, 5days/w,<br>4w(20h) | FMA | upper-limb |
| 14 | 2020 | Chew et al | 19(10/9) | 52.2±11.8/<br>56.4±9.6 | 31.3±24.5/<br>33.3±15.1 | tDCS-BCI to drive<br>MIT Manus robot | Sham tDCS-BCI | 5days/w, 2w | FMA | upper-limb |
| 15 | 2017 | Hong et al | 18(9/9) | 52.8±12.3/<br>56.4±9.6 | 33.9±24.6/<br>33.3±15.1 | tDCS-BCI to drive<br>MIT Manus robot | Sham tDCS-BCI | 40mins/day, 5days/w,<br>2w(6.67h) | FMA | upper-limb |
| 16 | 2021 | Hu et al | 18(10/8) | 52.1±11.7/<br>54.6±8.5 | 35.07±24.03/<br>35.87±15.53 | tDCS-BCI to drive<br>MIT Manus robot | Sham tDCS-BCI | 40mins/days, 5days/w,<br>2w | FMA | upper-limb |
| 17 | 2019 | Mane et al | 19(10/9) | 52.1±11.6/<br>56.3±9.5 | 35.1±24.1/<br>34.0±15.5 | tDCS-BCI to drive<br>MIT Manus robot | Sham tDCS-BCI | 60 mins/day, 10days (10h) | FMA | upper-limb |
| 18 | 2018 | Biasiucci | 27(14/13) | 56.36±9.91/<br>59.00±12.36 | 39.79±45.90/<br>33.46±30.51 | BCI to triggered FES | Sham-FES | 60mins/day, 2days/w,<br>5w(10h) | FMA, MRC | upper-limb |
| 19 | 2021 | Chen et al | 32(16/16) | 59.31/<br>57.93 | 6.27/<br>5.73 | BCI to trigger FES | Sham-FES | 40 mins/time, 4 times/w,<br>11times (29.3h) | FMA, MMT | upper-limb |
| 20 | 2016 | Jang et al | 20(10/10) | 61.10±13.77/<br>61.70±12.09 | 4.40±0.97/<br>4.10±0.74 | BCI to trigger FES | FES | 20 mins/days, 5days/w,<br>6w (10h) | MFT | upper-limb |
| 21 | 2016 | Kim et al | 30(15/15) | 59.07±8.07/<br>59.93±9.79 | 8.27±1.98/<br>7.80±1.78 | AOT-BCI to trigger<br>FES | Conventional<br>therapy | 30 mins/day, 5 days/w,<br>4w(10h) | FMA, MAL,<br>MBI, ROM | upper-limb |
| 22 | 2020 | Lee et al | 26(13/13) | 55.15±11.57/<br>58.30±9.19 | 7.46±1.61/<br>8.30±1.97 | AOT-BCI to trigger<br>FES | Conventional<br>therapy | 30 mins/days, 5days/w,<br>4w(10h) | FMA,<br>WMFT,<br>MAL, MBI | upper-limb |
| 23 | 2016 | Leeb et al | 18(9/9) | 55.1±11.0 | 37.3±43.9 | BCI to trigger FES | Sham-BCI | 60 mins/day, 2day/w,<br>5w(10h) | FMA | upper-limb |
| 24 | 2013 | Li et al | 14(7/7) | 66.3±4.53/<br>67.1±5.51 | 2.21±1.69/<br>2.79±1.85 | BCI to trigger FES | FES | 60 mins/day, 3days/w,<br>8w(24h) | FMA, ARAT | upper-limb |
| 25 | 2020 | Miao et al | 16(8/8) | 48.75±16.72/<br>50.25±17.11 | 18.25±10.93/<br>11.13±4.97 | BCI to trigger FES | Routine<br>rehabilitation | 3 sessions/w,<br>4w(12sessions) | FMA | upper-limb |
| 26 | 2021 | Hu et al | 12(7/5) | 44.9±7.5/<br>60.4±16.8 | 7.9±6.5/<br>7.3±4.5 | BCI with sensory and<br>virtual feedback | Classic MI | 30mins/day | FMA, ARAT,<br>MSS, BI | upper-limb |
| 27 | 2013 | Mihara et al | 20(10/10) | 58.1±8.3 | 4.5±1.27 | BCI(fNIRS) with visual feedback | Sham-BCI | 60 mins/days, 3days/w, 2w (6h) | FMA, ARAT, MAL, KVIQ-10 | upper-limb |
| 28 | 2015 | Pichiorri et al | 28(14/14) | 64.1±8.4/<br>59.6±12.7 | 2.7±1.7/<br>2.5±1.2 | BCI with visual feedback | Classic MI | 60 mins/day, 3days/w, 4w(12h) | FMA, MRC, NIHSS | upper-limb |
| 29 | 2015 | Chung et al | 10(5/5) | 43.6/<br>50.2 | 16.4±?/<br>8.4±? | BCI to trigger FES | FES | 30 mins/day, 5days/w, 1w(2.5h) | Time-up and go test | lower-limb |
| 30 | 2020 | Chung et al | 25(13/12) | 52.0±14.6/<br>54.1±14.7 | 11.3±5.6/<br>16.3±7.3 | BCI to triggered FES | FES | 30 mins/day, 3 days/w, 5w(7.5h) | Time-up and go test, BBS | lower-limb |
| 31 | 2021 | Li et al | 25(12/13) | 51.24±7.80/<br>49.56±8.40 | 1.07±0.35/<br>1.13±0.30 | BCI to drive rehabilitation robot | Pedal training | 30 mins/day, 5 days/w, 4w(10h) | FMA, NIHSS | lower-limb |
| 32 | 2016 | Mrachacz-Kersting et al | 22(13/9) | 46.31±12.51/<br>55.22±10.63 | 15.38±6.2/<br>18.00±4.47 | BCI to trigger FES | Sham-FES | 5-10mins/day, 3days/w, 1w(15-30mins) | FMA, 10-m walk test, ASS | lower-limb |
| 33 | 2018 | Mrachacz-Kersting et al | 24(12/12) | 60.8±7.9/<br>61.5±8.9 | 1.72±1.02/<br>1.88±1.16 | BCI to trigger FES | Sham-FES | 25mins/day, 3days/w, 4w(12h) | FMA, ASS, FAC, 10m walk test | lower-limb |
| 34 | 2021 | Yuan et al | 30(16/14) | / | Subacute stroke | BCI to drive robot | Pedal training | 2sessions/day, 6days/w, 2w(24 sessions) | cFMA | lower-limb |
| 35 | 2022 | Zhao et al | 28(14/14) | 50.1±11.1/<br>56.1±11.5 | 1.57±1.77/<br>0.92±0.6 | SSVEP-BCI to drive robot | Sham BCI | 30mins/day, 6days/w, 4w(12h) | FMA | lower-limb |

BCI indicates brain computer interface; BMI, brain machine interface; Exp, experimental group; Cont, control group; MI, motor imaginary; tDCS, transcranial direct current stimulation; FES, functional electronic stimulation; FMA, fugl-meyer assessment; cFMA, modified FMA; ADL, activity daily living; GAS, goal attainment scale; MAL, motor activity log; MAS, modified ashworth scale; MBI, modified barthel index; MFT, motor function test; MRC, medical research council scale; MMT, manual muscle testing; MSS, motor status scale; BI, barthel index; ROM, range of motion; KVIQ, kinesthetic and visual imagery questionnaire; BBS, berg balance scale; ASS, ashworth scale for spasticity; ARAT, action research arm test; NIHSS, national institute of health stroke scale; SSVEP, steady-state visually evoked potentials; WMFT, wolf motor function test.

### Methodological quality

Two authors independently assessed the methodological quality and the risk of bias of the included studies, any disagreement was resolved by discussion. The results of this evaluation are illustrated in **Supplementary Materials Figure S1 - S2**, which presented the assessment for each risk of bias item for each included study and the percentages across all included studies. Most of the studies were double-blind or single-blind and nearly half of the studies provided the method of randomization and allocation concealment.

### Adverse effects

Most of the included studies didn’t report any adverse effect during trials. Ten of the studies reported that no serious adverse effects were found in the participants during and after BCI training or intervention s [13, 14, 24, 28, 31, 38, 42, 45, 50, 52]. However, in these studies, a few participants did experience transient mild adverse reactions, such as hemiplegic shoulder pain [14], fatigue [14, 38], nausea [14], headache [14], dizziness [38, 42], and allergic to electrode pads [13]. Overall, the adverse effects were minimal and the BCI interventions were well-tolerated.

### Meta-analysis

#### Overall effect on motor function

The motor function of the patients with stroke was assessed by FMA in 30 out of 36 studies, involving 661 stroke patients. The pooled analysis of these studies showed a significant effect on the improvement of motor function with BCI, as compared with control interventions (SMD = 0.56, 95% CI = 0.25-0.81, *p* = 0.0002; I^2^ = 66%) (**Table 2, Figure 2(A)**). No evidence of publication bias was noted according to the Egger’s test conducted (t = 0.37, *p* = 0.711). The funnel plot looked generally symmetrical (**Figure 2(B)**).

**Figure 2.**
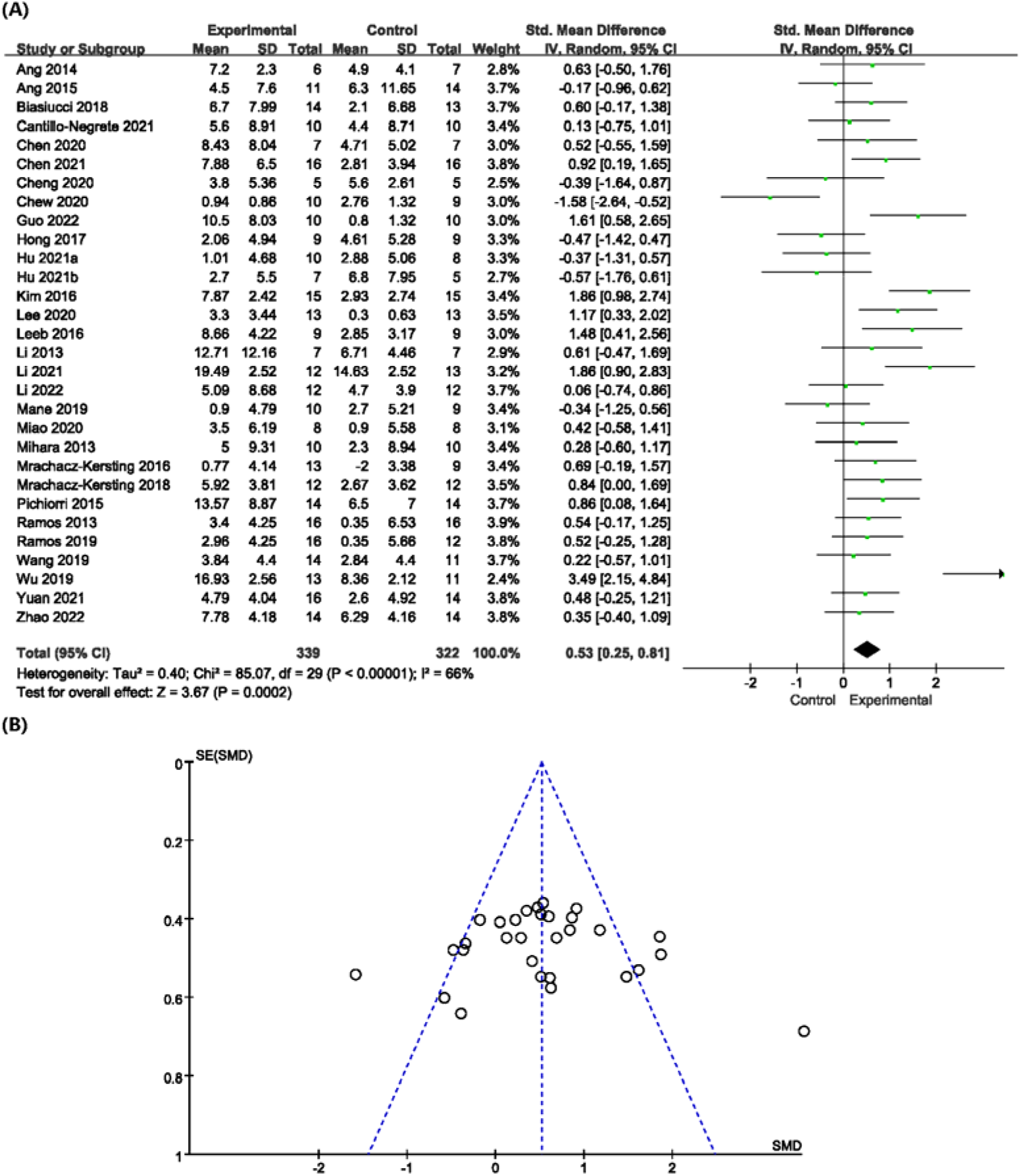
(A) Comparison of the therapeutic effects of BCI intervention and control intervention on motor function of stroke patients. The results revealed significant overall effects on improving motor recovery (SMD = 0.56, 95% CI = 0.25-0.81, p = 0.0002). (B) The funnel plot looked generally symmetrical. Egger’s test didn’t show significant publication bias (t = 0.37, p = 0.711).

**Table 2.**
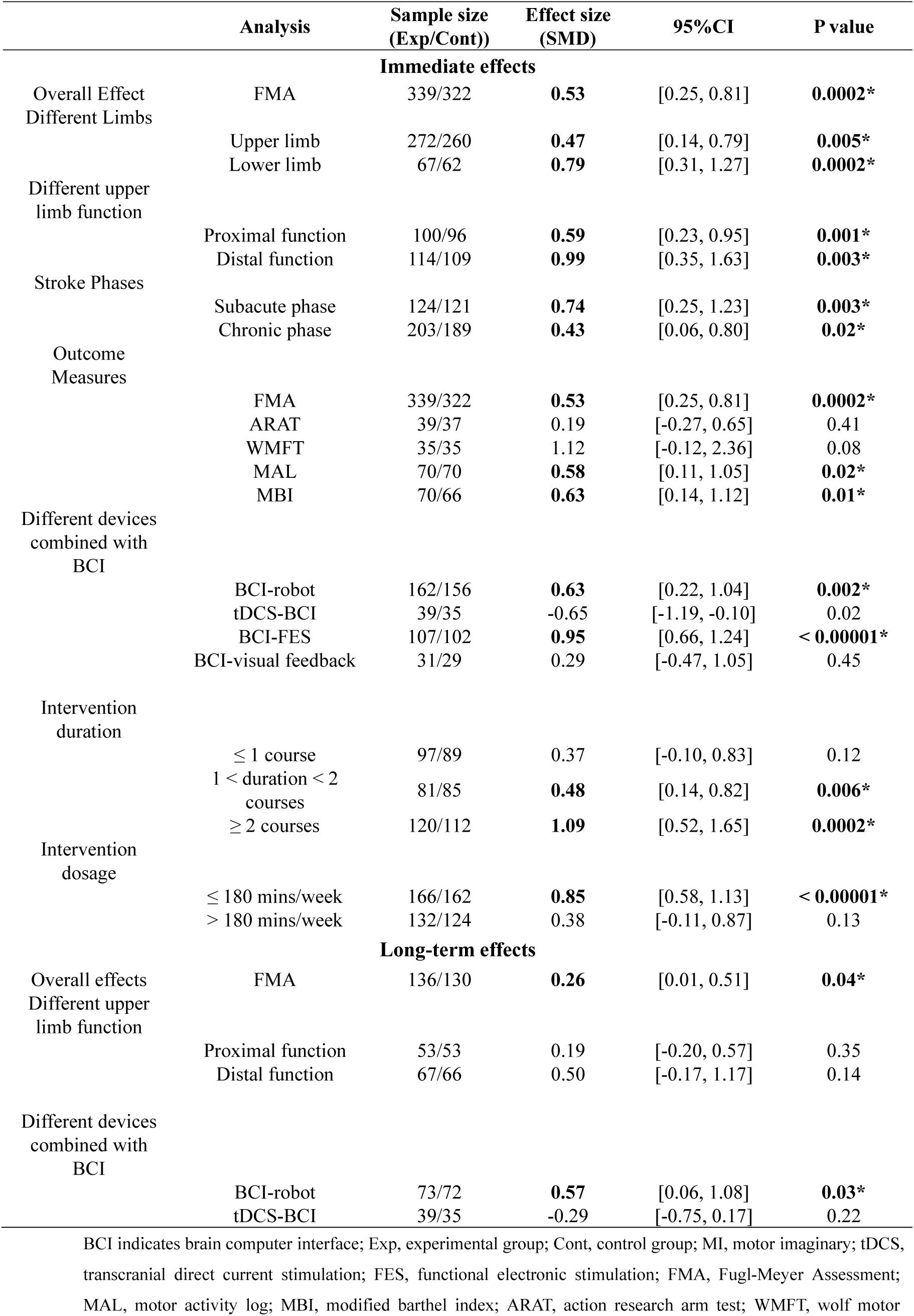
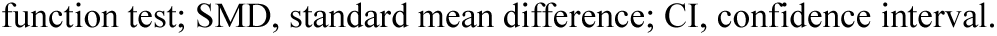
Summary of the effect size of BCI.

|  | Analysis | Sample size<br>(Exp/Cont)) | Effect size<br>(SMD) | 95%CI | P value |
| --- | --- | --- | --- | --- | --- |
| **Immediate effects** |  |  |  |  |  |
| Overall Effect<br>Different Limbs | FMA | 339/322 | **0.53** | [0.25, 0.81] | **0.0002\*** |
|  | Upper limb | 272/260 | **0.47** | [0.14, 0.79] | **0.005\*** |
|  | Lower limb | 67/62 | **0.79** | [0.31, 1.27] | **0.0002\*** |
| Different upper<br>limb function | Proximal function | 100/96 | **0.59** | [0.23, 0.95] | **0.001\*** |
|  | Distal function | 114/109 | **0.99** | [0.35, 1.63] | **0.003\*** |
| Stroke Phases | Subacute phase | 124/121 | **0.74** | [0.25, 1.23] | **0.003\*** |
|  | Chronic phase | 203/189 | **0.43** | [0.06, 0.80] | **0.02\*** |
| Outcome<br>Measures | FMA | 339/322 | **0.53** | [0.25, 0.81] | **0.0002\*** |
|  | ARAT | 39/37 | 0.19 | [-0.27, 0.65] | 0.41 |
|  | WMFT | 35/35 | 1.12 | [-0.12, 2.36] | 0.08 |
|  | MAL | 70/70 | **0.58** | [0.11, 1.05] | **0.02\*** |
|  | MBI | 70/66 | **0.63** | [0.14, 1.12] | **0.01\*** |
| Different devices<br>combined with<br>BCI | BCI-robot | 162/156 | **0.63** | [0.22, 1.04] | **0.002\*** |
|  | tDCS-BCI | 39/35 | -0.65 | [-1.19, -0.10] | 0.02 |
|  | BCI-FES | 107/102 | **0.95** | [0.66, 1.24] | **< 0.00001\*** |
|  | BCI-visual feedback | 31/29 | 0.29 | [-0.47, 1.05] | 0.45 |
| Intervention<br>duration | ≤ 1 course | 97/89 | 0.37 | [-0.10, 0.83] | 0.12 |
|  | 1 < duration < 2<br>courses | 81/85 | **0.48** | [0.14, 0.82] | **0.006\*** |
|  | ≥ 2 courses | 120/112 | **1.09** | [0.52, 1.65] | **0.0002\*** |
| Intervention<br>dosage | ≤ 180 mins/week | 166/162 | **0.85** | [0.58, 1.13] | **< 0.00001\*** |
|  | > 180 mins/week | 132/124 | 0.38 | [-0.11, 0.87] | 0.13 |
| **Long-term effects** |  |  |  |  |  |
| Overall effects<br>Different upper<br>limb function | FMA | 136/130 | **0.26** | [0.01, 0.51] | **0.04\*** |
|  | Proximal function | 53/53 | 0.19 | [-0.20, 0.57] | 0.35 |
|  | Distal function | 67/66 | 0.50 | [-0.17, 1.17] | 0.14 |
| Different devices<br>combined with<br>BCI | BCI-robot | 73/72 | **0.57** | [0.06, 1.08] | **0.03\*** |
|  | tDCS-BCI | 39/35 | -0.29 | [-0.75, 0.17] | 0.22 |

BCI indicates brain computer interface; Exp, experimental group; Cont, control group; MI, motor imaginary; tDCS, transcranial direct current stimulation; FES, functional electronic stimulation; FMA, Fugl-Meyer Assessment; MAL, motor activity log; MBI, modified barthel index; ARAT, action research arm test; WMFT, wolf motor
function test; SMD, standard mean difference; CI, confidence interval.

#### Effects on upper-limb versus lower-limb

Twenty-five and five studies reported the motor function as measured by FMA of upper-limb and lower-limb of stroke patients, respectively. Subgroup analysis based on the limb of treatment revealed that BCI had significant effects on both upper-limb (SMD = 0.47, 95% CI = 0.14-0.79, *p* = 0.005; I^2^ = 69%) and lower-limb (SMD = 0.79, 95% CI = 0.31-1.27, *p* = 0.001; I^2^ = 42%) relative to control intervention. Lower-limb tended to benefit more positive therapeutic effect from BCI (**Table 2, Figure 3**).

**Figure 3.**
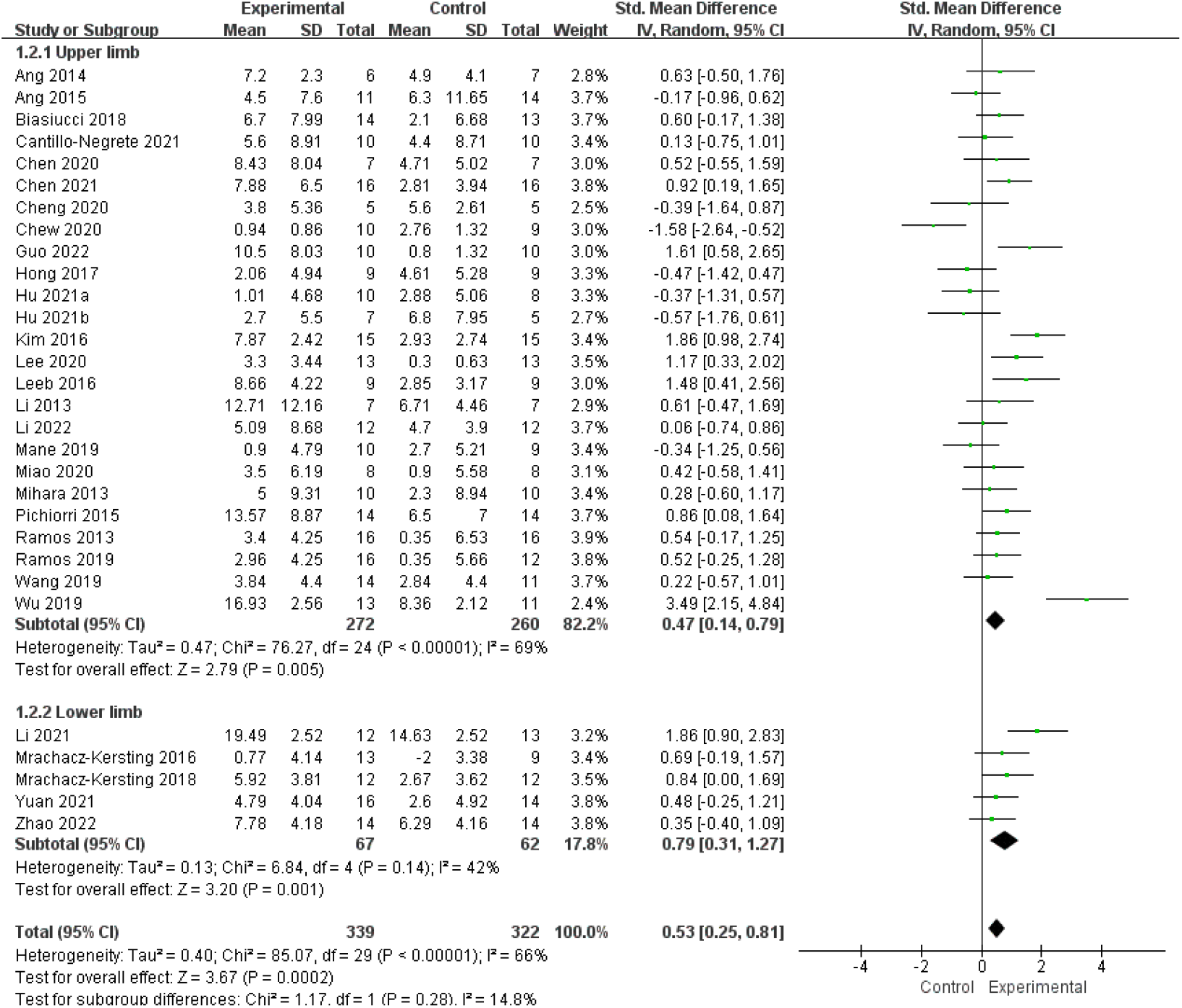
A subgroup analysis of the effects of different limbs. The results revealed that BCI had significant effects on both upper-limb (SMD = 0.47, 95% CI = 0.14-0.79, *p* = 0.005) and lower-limb (SMD = 0.79, 95% CI = 0.31-1.27, *p* = 0.001).

#### Effects on proximal versus distal function of upper-limb

To further clarify the effect of BCI on proximal and distal part of the motor functions of upper-limb, a subgroup analysis based on different functions of upper-limb was conducted. The pooled results showed greater mean effect size for distal functions (SMD = 0.99, 95% CI = 0.35-1.63, *p* = 0.003; I^2^ = 79%) than proximal functions of upper-limb (SMD = 0.59, 95% CI = 0.23-0.95, *p* = 0.001; I^2^ = 32%) (**Table 2, Figure 4**).

**Figure 4.**
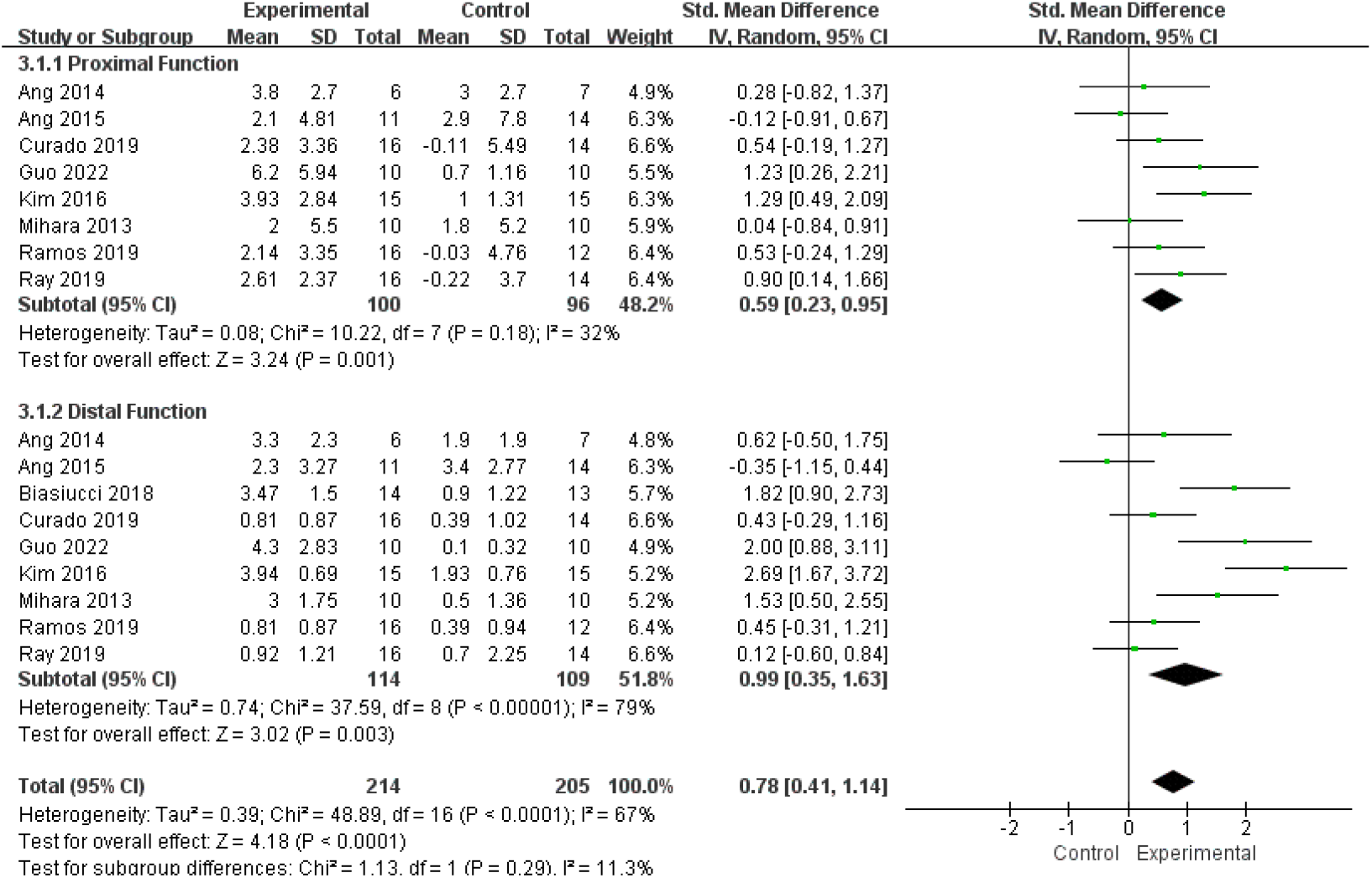
A subgroup analysis of the effects of different functions of upper-limb. The pooled results showed greater mean effect size for distal functions (SMD = 0.99, 95% CI = 0.35-1.63, p = 0.003) than proximal functions of upper-limb (SMD = 0.59, 95% CI = 0.23-0.95, p = 0.001).

#### Effects on subacute versus chronic stroke patients

Eleven studies recruited patients in the subacute phase and 19 studies focused on the chronic stroke patients. The mean effect size of subacute stroke patients (SMD = 0.74, 95% CI = 0.25-1.23, *p* = 0.003; I^2^ = 69%) was higher than that of chronic stroke patients (SMD = 0.43, 95% CI = 0.06-0.80, *p* = 0.02; I^2^ = 66%). This difference, however, was not significant (**Table 2, Supplementary Materials Figure S3**).

#### Effects of BCI combined with different devices

In the included studies, the upper/lower extremity robot, arm orthosis, tDCS, FES and visual feedback were used to combine with BCI. A subgroup analysis based on different devices combined with BCI indicated that BCI-based robot training (SMD = 0.63, 95% CI = 0.25-1.04, *p* = 0.002; I^2^ = 66%) and FES triggered by BCI (SMD = 0.95, 95% CI = 0.66-1.24, *p* < 10^-3^; I^2^ = 0%) had significant positive effect on motor function recovery, compared with control intervention. However, no significant effect size was observed for BCI training alone (SMD = 0.29, 95% CI = −0.47-1.05, *p* = 0.45; I^2^ = 50%) and BCI training combined with tDCS (SMD = −0.65, 95% CI = −1.19 - −0.1, *p* = 0.02; I^2^ = 22%) (**Table 2, Figure 5**).

**Figure 5.**
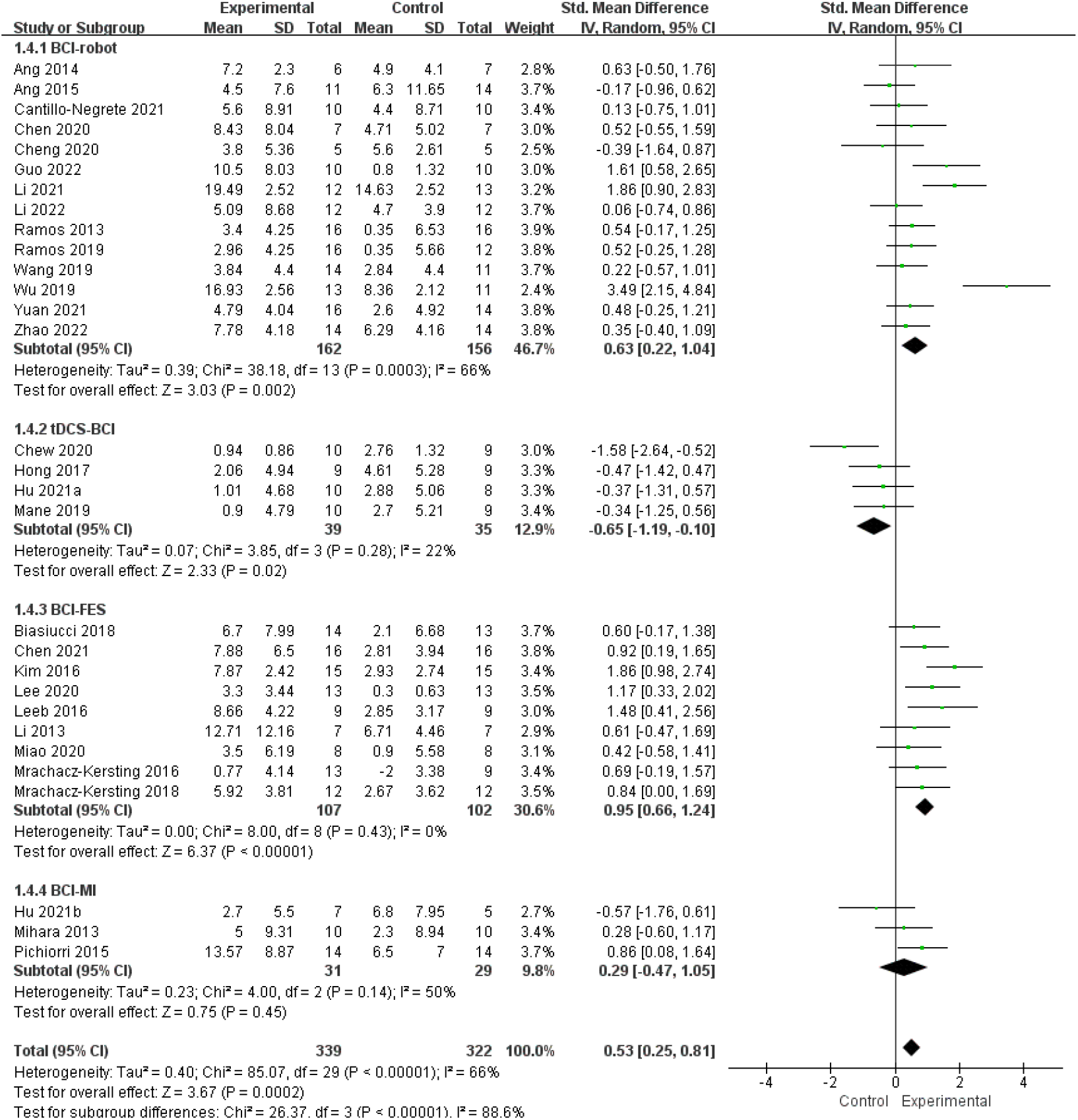
A subgroup analysis of the effects of different devices combined with BCI. The results revealed that only FES triggered by BCI (SMD = 0.95, 95% CI = 0.66-1.24, *p* < 10^-3^) and BCI-based robot training (SMD = 0.63, 95% CI = 0.25-1.04, *p* = 0.002) showed significant effects on motor recovery. No significant effect size was observed for BCI training alone (SMD = 0.29, 95% CI = −0.47-1.05, *p* = 0.45) and BCI training combined with tDCS (SMD = −0.65, 95% CI = −1.19 - −0.1, *p* = 0.02).

#### Comparison of different outcome measures

In addition to FMA, several other assessment scales for motor function were also used in these studies. Among these, ARAT, WMFT, MAL, and MBI were used to evaluate different aspects of motor function. A subgroup analysis based on these scales observed that BCI had significant mean effect sizes for MAL (SMD = 0.58, 95% CI = 0.11-1.05, *p* = 0.02; I^2^ =47%) and MBI (SMD = 0.63, 95% CI = 0.14-1.12, *p* = 0.01; I^2^ =48%). No significant mean effect size was found for ARAT (SMD = 0.19, 95% CI = −0.27-0.65, *p* = 0.41; I^2^ =0%) and WMFT (SMD = 1.12, 95% CI = −0.12-2.36, *p* = 0.08; I^2^ =82%) (**Table 2, Supplementary Materials Figure S4**).

#### Effects of the intervention duration and dosage

The duration of rehabilitation is the most important factor for the therapeutic effect. Generally, a 10-day of intervention is considered as a course of rehabilitation of stroke patients. Therefore, these studies were divided to three groups: ≤ 1 course; 1 - 2 courses; and ≥ 2 courses. The mean effect sizes of the subgroup analysis for the three groups were (SMD = 0.37, 95% CI = −0.10-0.83, *p* = 0.12; I^2^ =58%), (SMD = 0.64, 95% CI = 0.14-0.82, *p* = 0.006; I^2^ =14%), and (SMD = 1.09, 95% CI = 0.52-1.65, *p* = 0.0002; I^2^ =74%), respectively. The pooled results indicated that longer BCI intervention led to better improvement on motor function (**Table 2, Supplementary Materials Figure S5**).

In addition, the time of treatment per week also have a significant influence on the motor recovery. Counterintuitively, the subgroup analysis showed that <180 minutes of BCI per week dosage (SMD = 0.85, 95% CI = 0.58-1.13, *p* < 10^-3^; I^2^ =29%) has a more significant mean effect size than dosage of >180 minutes (SMD = 0.38, 95% CI = −0.11-0.87, *p* = 0.13; I^2^ =71%) BCI per week (**Table 2, Supplementary Materials Figure S6**).

#### Long-term effects on motor function

To clarify long-term sustained effects on the recovery of motor function of stroke patients, the data of FMA in long-term follow-ups were extracted. Significant mean effect size was detected for BCI (SMD = 0.26, 95% CI = 0.01-0.51, *p* = 0.04; I^2^ =45%), compared to control intervention (**Table 2, Supplementary Materials Figure S7**) Further subgroup analysis based on BCI strategies observed significant sustained mean effect size for BCI-based robot training (SMD = 0.57, 95% CI = 0.06-1.08, *p* = 0.03; I^2^ =53%) (**Table 2, Supplementary Materials Figure S8**). No significant effect size was found for other BCI strategies.

## Discussion

Although a number of studies have investigated the efficacy of BCI system on motor rehabilitation in stroke patients, information about differential effects on upper-limb and lower-limb, gross and fine motor function of upper-limb, therapeutic dosages, different BCI strategies, and long-term effects have not been synthesized. It is of importance to conduct quantitative estimates of the detailed efficacy of different body part, specific functions, durations and dosages of BCI training for helping determine the clinical therapeutic plan. For the current meta-analysis, we conducted a comprehensive analysis and obtained as much as evidence as possible from the literature on the effect of BCI on the motor recovery of stroke patients. We noted that lower-limb obtained more positive effects than upper-limb from BCI. The subgroup analysis indicated a greater intervention effect for distal function than proximal function of upper-limb, and the BCI combined with FES was more effective than BCI combined with robot, tDCS, or visual feedback devices. Furthermore, higher significant effect size was found for the longer durations of overall BCI training, and robots’ long-term effect. However, more is not always better for the therapeutic dosages, as the results show dosage of >180 min of training per week did not lead to better clinical outcome than dosage of ≤180 min of training per week.

### Different body parts obtained distinct effect from BCI

All available data of RCT studies on both upper-limb and lower-limb were applied to evaluate the overall effect of BCI training on the motor recovery of stroke patients. The pooled analysis showed significant overall effect size (SMD = 0.53, *p* = 0.0002) and greater effect size on lower-limb (SMD = 0.79, *p* = 0.001) than upper-limb (SMD = 0.47, *p* = 0.005). The similar significant effects of BCI intervention for the paretic upper-limb of stroke patients has been reported and demonstrated in several previous meta-analyses [16, 18, 19]. Therefore, we conclude that stroke patients can benefit from BCI intervention, in terms of improving the motor recovery of the hemiparetic upper extremity. Our updated study included more trials and larger sample size, providing further robust evidence of the positive effect of BCI on upper-limb. In addition, we provide the first pooled analysis evaluating the effectiveness of BCI on lower extremity function. The comparison of the effect sizes between upper-limb and lower-limb indicated that the lower-limb functions seemed to respond better to BCI training than upper-limb functions, which is consistent with a previous meta-analysis on the effects of tDCS in stroke patients [61] and a previous systemic review on MI training, in which higher effect sizes on both balance and gait were found than upper-limb [62]. This difference could be explained by the complexity of the function between upper-limb and lower-limb: lower-limb functions were simpler than upper-limb functions, and require less neural resources. Consequently, they respond faster than upper-limb functions when presented similar therapy. Furthermore, imaging study has demonstrated that the impairment of the microstructural integrity of the ipsilesional corticospinal tract induced by stroke may affect more on the upper-limb than lower-limb [63]. However, relative to the number of upper-limb studies, only a limited number of lower trials were included in out meta-analysis. The greater effect size of lower-limb obtained in our study needs to be further clarified in the future studies with more studies and larger sample size.

### BCI induced distal to proximal motor recovery

For the upper-limb, more prominent therapeutic effect on the distal limb function (SMD = 0.99) was observed than the proximal limb function (SMD = 0.59) in our study. Similar results have been reported in a meta-analysis that evaluated the effect of transcranial magnetic stimulation (rTMS) on stroke-induced upper-limb motor deficit [64]. More detailed information was shown in this study which revealed that the effectiveness of the rTMS appears to follow a descending order of finger, hand strength, activity dexterity, and body function. In addition, both the robotic rehabilitation [65] and EMG-driven NMES-robotic support training system [66] were also found more effective to the distal function that the proximal parts in the whole upper extremity. Another study provide evidence for this phenomenon that following a robot-assisted rehabilitation treatment there is a distal-to-proximal generalization in stroke patients [67]. Therefore, the possible reason for the consistent results is that robot-assisted rehabilitation driven by BCI was applied in 8 out of 9 included studies in our subgroup analysis of different parts of upper-limb. However, the mechanism is still unknown.

### Optimal duration and dosage of BCI

The optimal therapeutic dosage and duration are critical issues in the clinical diagnosis and treatment. In our study, the subgroup analysis among the cumulative duration and dosage of BCI training showed greater significant effects for long-term (> 10 days of treatment) and lower dosage rehabilitation (≤ 180 mins/week) respectively. Similarly, a previous meta-analysis on rTMS revealed that the increased efficacy of rTMS on antidepression therapy was associated with the increasing rTMS sessions. The best antidepressant effect of rTMS was realized when delivering 1200-1500 pulses/day rather than higher doses [68]. As a rehabilitation technology, previous studies have demonstrated that BCI could foster neuroplasticity and realize the cortical reorganization of the lesioned hemisphere through manipulation or self-regulation neurophysiological activity facilitating motor recovery [69]. Adequate time involvement is very important for the reorganization of the impaired cerebral cortex, which may be the explanation for the better effect of long-term BCI treatment. However, this reorganization should be realized under certain proper number of training each time. Overtreatment induced mental and physical fatigue may lead to negative effects on the neural activity and disturb the recovery of motor function. Therefore, long duration of BCI-based rehabilitative intervention is important but should be limited to an appropriate doses per week.

### Better devices combined with BCI and better stoke stage for intervention

Although the pooled subgroup analysis regarding the stroke phases, external devices combined with BCI, and corresponding long-term effects of BCI interventions have been documented in several previous reviews [16, 18, 19], the results were inconsistent at best and far from between. For the stroke onset, Nojima et al. reported the same effect size of BCI in the subacute and chronic stroke patients [19]. However, higher intervention effect was found for the subacute subgroup in the study of Cervera [16], which is more in agreement with our findings. On the other hand, patients in both subacute and chronic phase showed significant improvements in our study. Regarding to the comparison of external devices combined with BCI, the two studies observed the significantly superior clinical effects of BCI combined with FES than with other devices [18, 19]. They also found that the tDCS prior to the BCI intervention could not facilitate the effect of BCI for motor recovery [18]. All the result are consistent with our findings, except that we also found the significant immediate and long-lasting effect size when BCI was combined with robot. The possible reason for the discrepancy between studies should be the number of trials included in the pooled analysis: a greater number of trials were included in our updated meta-analysis (14 vs 5). It has been known that the clinical recovery generally occurs within 6 months after the onset of stroke, the earlier, the more rapid [70]. This could explain the higher intervention effects on subacute stroke patients. The earlier rehabilitation, the better recovery. The possible mechanism for the larger effect size of BCI combined with FES could be explained by the specific potentiating neural activation of the primary sensorimotor cortex after receiving somatosensory stimulation during motor tasks [71]. The essential role of somatosensory information in the production of high-quality motor outputs could realize a better recovery of motor function [72]. Further, the sensory input for the closed-loop BCI system from the external device could promote the self-regulation of mental activity and enhance brain plasticity induction [73]. In summary, the existing and the current analysis indicated that stroke patients in subacute phase may obtain more therapeutic effects from BCI training. BCI combined with robot could induce significant immediate and long-lasting therapeutic effects on motor recovery. BCI combined with FES may be a better choice than BCI combined with robot and other devices. Further studies on the long-term effect BCI combined with FES should be conducted in the future studies.

### Underlying Neural Mechanisms

Currently, almost all of the commonly used technologies in brain science research including EEG, functional magnetic resonance imaging (fMRI), diffusion tensor imaging (DTI), functional near-infrared spectroscopy (FNIRS), and transcranial magnetic stimulation (TMS) have been used to investigate the potential neural mechanisms of BCI on motor recovery. These studies indicated that BCI could induce physiological and structural neuroplasticity, *i.e.* to reorganization of the impaired cerebral cortex. These functional and structural reorganization reflected as the modulation on the functional spontaneous brain activity [33], evoked brain activity/excitability [43, 74, 75], microstructure [76, 77], interhemispheric imbalance [14, 33], and cortical-subcortical connectivity [78] of the motor related brain regions. In most if not all studies, such changes would lead to functional improvements for stroke patients. However, the results were not as conclusive as one would hope, due to small number of studies and discrepancy reported among them. Evidently, more studies are needed to further elucidate the exact mechanisms of recovery underlying BCI training and the factors influencing rehabilitation effects in stroke patients.

## Conclusions

This study provides evidence for the benefit of BCI training in stroke rehabilitation. It is safe, and provides significant immediate and long-lasting effects on the improvement of both upper-limb and lower-limb of stroke patients. Earlier intervention may obtain more positive effects, especially for the distal function of upper-limb. BCI combined with FES seems to be more effective than BCI combined with robot and other external devices. However, BCI combined with robot provide evidence for the long-term effects. The applying tDCS prior to the BCI intervention does not seem to ‘prime’ the cortex with any significant effect. To achieve a better therapeutic effect, at least 10 days of BCI training should be provided. The weekly dosage may need to be controlled to ≤ 180min. More studies are also required to clarify the reliability of these results due to a substantial heterogeneity in this meta-analysis.

## Data Availability

Data available on request from the authors.

## Conflict of interest

The authors declared no potential conflict of interest with respect to the research, authorship, and publication of this article.

## Funding

This work was supported by the National Natural Science Foundation of China (NSFC, No. 82172540).

## Supplementary Material

Data extracted from included studies in this meta-analysis, data used for all analyses and analytic code can be made available upon request.

Figure S1 - S8

## Notes

### Competing Interest Statement

The authors have declared no competing interest.

